# Low back pain and telecommuting in Japan: influence of work environment quality

**DOI:** 10.1101/2022.01.27.22269946

**Authors:** Ryutaro Matsugaki, Tomohiro Ishimaru, Ayako Hino, Keiji Muramatsu, Tomohisa Nagata, Kazunori Ikegami, Seiichiro Tateishi, Mayumi Tsuji, Shinya Matsuda, Yoshihisa Fujino, the CORoNaWork Project

**Author notes:** **Corresponding author:** Yoshihisa Fujino, M.D., M.P.H., Ph.D., Department of Environmental Epidemiology, Institute of Industrial Ecological Sciences, University of Occupational and Environmental Health, Japan, 1-1, Iseigaoka, Yahatanishiku, Kitakyushu, 807-8555, Japan. **Author contributions:** R.M. wrote the manuscript; T.I., A.H., K.M., T.N., K.I., S.T., M.T., and S.M. reviewed the manuscript, provided advice on interpretation, and secured funding for research; Y.F. planned the overall survey, created the questionnaire, analyzed the data, and drafted the manuscript.

## Abstract

**Objectives:** This study examined the relationship between frequency of working from home and low back pain (LBP), considering the quality of work environment.

**Methods:** The study was based on a cross-sectional internet-based survey. Of 33,302 respondents, data from 12,774 desk workers were retained for analysis. We used a 0–10 numerical rating scale to assess LBP. Work environment was assessed using five subjective questions. Mixed-effects logistic regression nested by city level was used to analyze the relationship between frequency of working from home and LBP, stratified by work environment condition.

**Results:** The prevalence of LBP was 21.0%. Among those reporting a poor work environment, as opposed to almost never working from home, the multivariate odds ratio (OR) of LBP were as follows: working from home less than 1 day per week: OR=1.25, 95% CI: 0.89–1.76, p=0.190); 2-3 days per week: OR=1.58, 95% CI: 1.16–2.16, p=0.004; and 4 or more days per week: OR=1.82, 95% CI: 1.38–2.40, p<0.001. By contrast, among those reporting a good work environment the OR of LBP did not increase as the frequency of working from home increased.

**Conclusions:** The relationship between LBP and frequency of working from home was found to vary with the quality of the work environment; more specifically, LBP was associated with frequency of teleworking in a poor work environment. This study suggests that employers should give more support their employees in promoting a good work environment to prevent LBP. (Words: 240/250)

## Introduction

Working from home (a form of “teleworking” or “telecommuting”) has been recommended worldwide since the outbreak of the coronavirus disease 2019 (COVID-19).^1^ For example the Japanese government has recommended work from home to counter the spread of COVID-19^2, 3^, and the percentage of telework including work from home increased from 15.4% to 22.5% between October 2019 and November 2020^4^. Work from home is expected to become an increasingly common way of working in the future, but impacts of work from home on health are still largely unknown.

Currently there is no consensus regarding the potential relationship between telework and lower back pain (LBP). A few recent studies have reported that work from home is associated with LBP^5-7^. Yoshimoto et al. reported an increased risk of LBP among workers who began teleworking and those who increased their frequency of telework during the COVID-19 pandemic^5^. Another report showed an association between the frequency of telework and the prevalence of LBP^7^. However, one report suggests that telework is not associated with exacerbation of LBP^8^, while another study reported a decrease in musculoskeletal pain including LBP among workers who teleworked in confinement due to the COVID-19 virus^9^.

Work from home and LBP may be associated with the quality of work environment. Previous studies have demonstrated such a relationship in office workers^10-13^. We reported that inadequate work environments (insufficient room to concentrate, and inadequate lighting, desk space and foot space) were associated with LBP among workers who work from home^14^. However, research on the relationship between the frequency of work from home and LBP that includes consideration of the quality of the work environment has yet to be conducted.

This study set out to examine the relationship between working from home and LBP, considering the quality of the work environment. Our hypothesis was that work from home and LBP are related when the work environment is poor, whereas they are not related when the latter is good.

## Methods

### Study Design and Subjects

We conducted a cross-sectional internet-based survey between December 22 and December 26, 2020. More details about the survey protocol are available in another article^15^. The survey targeted people currently in possession of an employment contract. The exclusion criteria were (1) giving identifiably false responses, (2) being not mainly a desk worker, and (3) working less than 5 days per week. Of the 33,302 respondents, 12,774 were retained for the final analysis.

This study respected the principles of the Declaration of Helsinki, and received approval by the ethics committee of the University of Occupational and Environmental Health, Japan (reference No. R2-079 and R3-006). Participants gave informed consent online through the website.

### Assessment of LBP

As in our previous study^14^, participants’ experience of LBP was assessed by their responses to two simple questions. The first question: “Have you experienced stiff shoulders or LBP in the past two weeks?” required a “yes” or “no” answer. If the participant answered “yes”, they continued to the following question, which focused on severity of LBP: “What was your average level of LBP in the past two weeks? (Please rate your pain from 0 to 10, where 0=no pain at all and 10=the most intense pain you have experienced).” We defined prevalent LBP as a pain intensity of 6 or higher, based on previous studies^16^.

### Assessment of telecommuting

Telecommuting status was assessed using the same question as in the previous study^14^: “Do you work at home? Please choose the answer that is closest to your current situation.” The respondents chose one of the five following options: 4 days a week or more, 2 to 3 days a week, 1 day a week, more than once a month but less than once a week, and almost never. The answers “1 day a week” and “more than once a month but less than once a week” were classified together as “less than once a week”.

### Assessment of telecommuting environment

We used the following five items to categorize telecommuting environments: 1) “Do you have a place or room where you can concentrate on your work?” 2) “Is your desk well-enough lit for you to work?” 3) “Do you have enough space on your desk to work?” 4) “Is there enough space to stretch your legs?” 5) “Are the temperature and humidity in the room where you work appropriate for working comfortably?” These five questions were known to be related to prevalence of LBP in telecommuting workers^14^. The respondents answered “yes” or “no” to each question. For the analysis, a telecommuting environment was classified as “good” if the number of yes answers to the five questions was 3 or more, and “poor” if the number of answers was 2 or fewer.

### Assessment of participant characteristics and other covariates

In the analysis we took into consideration the following socioeconomic factors: age, gender, body mass index (BMI), marital status, educational background (junior high school, high school, university or vocational school, junior college or technical college, or graduate school), and equivalent income (household income divided by the square root of the household size). Also included were the following lifestyle factors: smoking (currently smoking), drinking (consuming alcohol on two or more days per week), physical activity (walking or equivalent physical activities) for at least 1 h a day for more than 2 days a week).

To assess mental health status we used the Kessler 6 (K6)^17^, the Japanese version of which was validated previously^18^. This tool was developed to screen for mental disorders including depression and anxiety. It consists of six questions, with answers to each ranging from 0 (never) to 4 (always), according to frequency of experiencing the event described in the question within the past 30 days. The higher total score, the greater the potential for depression or anxiety disorder. We used a K6 score ≥5 as an indicator of the presence of psychological distress.

The company size (total number of people employed by the company where the participant works) was recorded in one of four categories, 1-9, 10-99, 100-999, and over 1,000.

### Statistical analysis

Age and BMI are expressed as continuous variables, using the mean and standard deviation. Other variables are presented categorically, using numerical values and percentages.

Mixed-effects logistic regression analysis nested by city level was conducted to analyze the relationship between telecommuting frequency and LBP. First, preliminary analysis confirmed a significant interaction between telecommuting frequency and telecommuting environment. Second, we estimated age-sex adjusted and multivariate adjusted odds ratios stratified by telecommuting environment. The multivariate model included age, sex, BMI, marital status, educational background, equivalent income, lifestyle habit, psychological distress, and company size.

All statistical analyses were performed with Stata software (Stata Statistical Software: Release 16; StataCorp LLC, TX, USA). Any p-values of <0.05 were considered statistically significant.

## Results

Table 1 presents details of participant characteristics. Of the 12,774 workers, 9,082 (71.1%) telecommuted almost never, 873 (6.8%) telecommuted less than 1 day per week, 953 (7.5%) 2 to 3 days a week, 1,866 (14.6%) telecommuted 4 days a week or more. Regarding the telecommuting environment, the percentage of those reporting “good” was 60.6% (5,505), 76.7% (670), 75.8% (722), and 84.5% (1,577) in groups that telecommuted almost never, less than 1 day, 2 to 3 days, and 4 days a week or more respectively. The overall prevalence of LBP was 21.0% (2,686/ 12,774 workers).

**Table 1.**
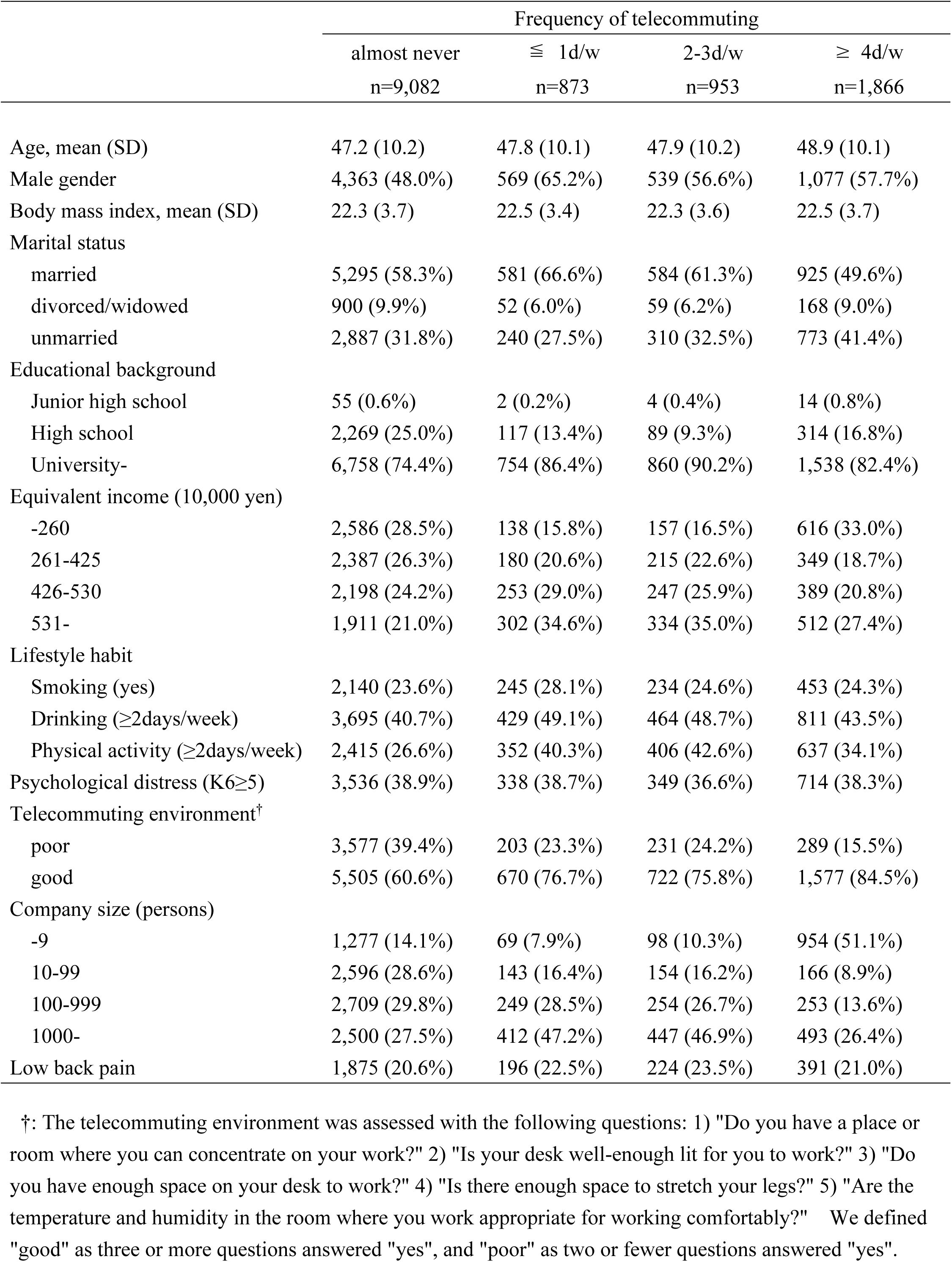
Participant characteristics

|  | Frequency of telecommuting |  |  |  |
| --- | --- | --- | --- | --- |
|  | almost never<br>n=9,082 | ≤ 1d/w<br>n=873 | 2-3d/w<br>n=953 | ≥ 4d/w<br>n=1,866 |
| Age, mean (SD) | 47.2 (10.2) | 47.8 (10.1) | 47.9 (10.2) | 48.9 (10.1) |
| Male gender | 4,363 (48.0%) | 569 (65.2%) | 539 (56.6%) | 1,077 (57.7%) |
| Body mass index, mean (SD) | 22.3 (3.7) | 22.5 (3.4) | 22.3 (3.6) | 22.5 (3.7) |
| Marital status |  |  |  |  |
| married | 5,295 (58.3%) | 581 (66.6%) | 584 (61.3%) | 925 (49.6%) |
| divorced/widowed | 900 (9.9%) | 52 (6.0%) | 59 (6.2%) | 168 (9.0%) |
| unmarried | 2,887 (31.8%) | 240 (27.5%) | 310 (32.5%) | 773 (41.4%) |
| Educational background |  |  |  |  |
| Junior high school | 55 (0.6%) | 2 (0.2%) | 4 (0.4%) | 14 (0.8%) |
| High school | 2,269 (25.0%) | 117 (13.4%) | 89 (9.3%) | 314 (16.8%) |
| University- | 6,758 (74.4%) | 754 (86.4%) | 860 (90.2%) | 1,538 (82.4%) |
| Equivalent income (10,000 yen) |  |  |  |  |
| -260 | 2,586 (28.5%) | 138 (15.8%) | 157 (16.5%) | 616 (33.0%) |
| 261-425 | 2,387 (26.3%) | 180 (20.6%) | 215 (22.6%) | 349 (18.7%) |
| 426-530 | 2,198 (24.2%) | 253 (29.0%) | 247 (25.9%) | 389 (20.8%) |
| 531- | 1,911 (21.0%) | 302 (34.6%) | 334 (35.0%) | 512 (27.4%) |
| Lifestyle habit |  |  |  |  |
| Smoking (yes) | 2,140 (23.6%) | 245 (28.1%) | 234 (24.6%) | 453 (24.3%) |
| Drinking (≥2days/week) | 3,695 (40.7%) | 429 (49.1%) | 464 (48.7%) | 811 (43.5%) |
| Physical activity (≥2days/week) | 2,415 (26.6%) | 352 (40.3%) | 406 (42.6%) | 637 (34.1%) |
| Psychological distress (K6≥5) | 3,536 (38.9%) | 338 (38.7%) | 349 (36.6%) | 714 (38.3%) |
| Telecommuting environment<sup>†</sup> |  |  |  |  |
| poor | 3,577 (39.4%) | 203 (23.3%) | 231 (24.2%) | 289 (15.5%) |
| good | 5,505 (60.6%) | 670 (76.7%) | 722 (75.8%) | 1,577 (84.5%) |
| Company size (persons) |  |  |  |  |
| -9 | 1,277 (14.1%) | 69 (7.9%) | 98 (10.3%) | 954 (51.1%) |
| 10-99 | 2,596 (28.6%) | 143 (16.4%) | 154 (16.2%) | 166 (8.9%) |
| 100-999 | 2,709 (29.8%) | 249 (28.5%) | 254 (26.7%) | 253 (13.6%) |
| 1000- | 2,500 (27.5%) | 412 (47.2%) | 447 (46.9%) | 493 (26.4%) |
| Low back pain | 1,875 (20.6%) | 196 (22.5%) | 224 (23.5%) | 391 (21.0%) |

<sup>†</sup>: The telecommuting environment was assessed with the following questions: 1) "Do you have a place or room where you can concentrate on your work?" 2) "Is your desk well-enough lit for you to work?" 3) "Do you have enough space on your desk to work?" 4) "Is there enough space to stretch your legs?" 5) "Are the temperature and humidity in the room where you work appropriate for working comfortably?" We defined "good" as three or more questions answered "yes", and "poor" as two or fewer questions answered "yes".

The odds ratios of LBP associated with telecommuting frequency stratified by telecommuting environment are shown in table 2. Among participants who had a good telecommuting environment, the odds ratios of LBP did not increase as telecommuting frequency increased. Outside of those who almost never telecommuted, the multivariate odds ratios of LBP in those who telecommuted less than 1 day per week, 2 to 3 days per week, and 4 days per week or more, were 1.15 (95% CI: 0.94–1.41, p=0.176), 1.17 (95% CI: 0.96–1.43, p=0.113), and 1.03 (95% CI: 0.88–1.21, p=0.705), respectively. By contrast, among those who had a poor telecommuting environment, the odds ratios of LBP did increase as telecommuting frequency increased. Again disregarding those who almost never telecommuted, the multivariate odds ratios of LBP were 1.25 (95% CI: 0.89–1.76, p=0.190), 1.58 (95% CI: 1.16–2.16, p=0.004), and 1.82 (95% CI: 1.38–2.40, p<0.001) for the less than 1 day per week group, 2 to 3 days a week, and 4 days a week or more groups, respectively.

**Table 2.** Odds ratio of low back pain associated with frequency of work from home stratified by work environment

|  | Age-sex adjusted |  |  |  | Multivariate† |  |  |  |
| --- | --- | --- | --- | --- | --- | --- | --- | --- |
|  | OR | 95%CI |  | p | OR | 95%CI |  | p |
| All participants (n=12,774) |  |  |  |  |  |  |  |  |
| Frequency of telecommuting |  |  |  |  |  |  |  |  |
| Almost never | Reference |  |  |  | Reference |  |  |  |
| ≦ 1d/w | 1.16 | 0.98 | 1.37 | 0.082 | 1.18 | 0.99 | 1.41 | 0.060 |
| 2-3d/w | 1.21 | 1.03 | 1.41 | 0.021 | 1.27 | 1.08 | 1.50 | 0.005 |
| ≥ 4d/w | 1.05 | 0.93 | 1.19 | 0.437 | 1.15 | 1.01 | 1.32 | 0.040 |
|  |  |  |  | 0.109* |  |  |  | 0.003* |
| Participants with a good telecommuting environment ‡ (n=8,474) |  |  |  |  |  |  |  |  |
| Frequency of telecommuting |  |  |  |  |  |  |  |  |
| Almost never | Reference |  |  |  | Reference |  |  |  |
| ≦ 1d/w | 1.13 | 0.92 | 1.37 | 0.238 | 1.15 | 0.94 | 1.41 | 0.176 |
| 2-3d/w | 1.11 | 0.92 | 1.34 | 0.282 | 1.17 | 0.96 | 1.43 | 0.113 |
| ≥ 4d/w | 0.94 | 0.82 | 1.09 | 0.431 | 1.03 | 0.88 | 1.21 | 0.705 |
|  |  |  |  | 0.754* |  |  |  | 0.340* |
| Participants with a poor telecommuting environment ‡ (n=4,300) |  |  |  |  |  |  |  |  |
| Frequency of telecommuting |  |  |  |  |  |  |  |  |
| Almost never | Reference |  |  |  | Reference |  |  |  |
| ≦ 1d/w | 1.31 | 0.94 | 1.81 | 0.111 | 1.25 | 0.89 | 1.76 | 0.190 |
| 2-3d/w | 1.57 | 1.17 | 2.11 | 0.003 | 1.58 | 1.16 | 2.16 | 0.004 |
| ≥ 4d/w | 1.80 | 1.39 | 2.34 | <0.001 | 1.82 | 1.38 | 2.40 | <0.001 |
|  |  |  |  | <0.001* |  |  |  | <0.001 |

†: The multivariate model adjusted for age, gender, body mass index, marital status, educational background, equivalent income, lifestyle habit (smoking, drinking, and physical activity), psychological status, and company size.
‡: The telecommuting environment was assessed with the following questions: 1) "Do you have a place or room where you can concentrate on your work?" 2) "Is your desk well-enough lit for you to work?" 3) "Do you have enough space on your desk to work?" ; 4) "Is there enough space to stretch your legs?" 5) "Are the temperature and humidity in the room where you work appropriate for working comfortably?"
We defined "good" as three or more questions answered "yes", and "poor" as two or fewer questions answered "yes".
\*p-value of trend

## Discussion

The relationship between LBP and work from home differed depending on the telecommuting environment. The results showed that there was no association between LBP and work from home when the work environment was good, whereas the prevalence of LBP increased with the frequency of work from home when the work environment was poor.

This study revealed that some people who work from home are doing so in poor work environments. While the survey was being conducted, the Japanese government responded to the rapid spread of COVID-19 by recommending the implementation of work from home to curb the spread of infection^2,3^. As a consequence, telecommuting may have been imposed involuntarily, regardless of the quality of the work environment. In a survey conducted contemporaneously with this survey, fewer than 50% of Japanese workers reported having a work desk and chair in their home that were suitable for work from home^4^. In this study, it was also shown that 15.5% of workers worked from home in a poor work environment even if they do so more than 4 times a week.

Our data revealed that the relationship between working from home and LBP was clearly different depending on whether the telecommuting practice was maintained or not. Tezuka et al. reported a relationship between frequency of work from home and physical symptoms including LBP in Japanese workers who started work from home under COVID-19^7^, but the dose-response relationship between LBP and frequency of work from home was not clear. Although that study did not focus on the participants’ work environment, it is possible that there was a mix of home workers with good and bad work environments. In this study, no dose-response relationship between frequency of work from home and LBP was clear when the work environment was not taken into consideration.

We found a dose-response relationship between the prevalence of LBP and the frequency of work from home when the work environment was poor. Awkward posture is known to be a risk factor for LBP^10, 13, 19^. Inadequate illumination of desks and inadequate space around desks can result in LBP by forcing workers into awkward postures. Cold temperatures are also a known risk factor for LBP^20-23^. Suboptimal temperature and humidity in rooms used for work from home can lead to LBP by increasing muscle tension in workers’ lower back. Furthermore, psychological stress is also considered to be a risk factor for LBP^24-26^, and the lack of a room dedicated to work from home may cause LBP in workers by increasing their psychological stress.

As the work environment of telecommuters is closely related to their personal life, it is more difficult than an office environment for employers to manage^14^. Therefore, employers need to provide workers who work from home with information and education on appropriate work environments so that they can manage themselves. For instance, the guidelines issued by the Ministry of Health, Labor and Welfare^27, 28^ may be useful, as they provide recommended telecommuting environment levels for lighting, temperature, humidity, etc.

This study has some limitations. First, the work environment was evaluated using subjective questions. Temperature and humidity could have been measured using a thermometer or hygrometer, and desk illumination could have been measured using an illuminance meter. However, there is currently no objective way to evaluate the space around a desk or a room where one can focus on work. Second, there is the problem of unmeasured confounds. It is known that factors such as position of the computer display^20^, sitting time^10^, and past symptoms^29^ affect LBP in workers, but these were not considered here. Whether these factors might have influenced our results is not clear. Finally, because of the study’s cross-sectional design, we cannot draw conclusions about any causal relationship. However, it seems unlikely that workers suffering from LBP would arrange the work environment in a way that would exacerbate their problem.

## Conclusion

We found that the relationship between LBP and work from home differed depending on the quality of the work environment. The evidence suggests that LBP is associated with work from home when the work environment is poor. Employers need to appreciate the importance of the telecommuting environment when asking employees to work from home. They should offer advice about appropriate work environments both when the environment for work from home has not yet been set up, and when it is already being used.

## Data Availability

For availability of data and material please contact the corresponding author.

## Acknowledgement

This study was supported and partly funded by the research grant from the University of Occupational and Environmental Health, Japan (no grant number); Japanese Ministry of Health, Labour and Welfare (H30-josei-ippan-002, H30-roudou-ippan-007, 19JA1004, 20JA1006, 210301-1, and 20HB1004); Anshin Zaidan (no grant number), the Collabo-Health Study Group (no grant number), and Hitachi Systems, Ltd. (no grant number) and scholarship donations from Chugai Pharmaceutical Co., Ltd. (no grant number). The funder was not involved in the study design, collection, analysis, interpretation of data, the writing of this article or the decision to submit it for publication. All authors declare no other competing interests.

The current members of the CORoNaWork Project, in alphabetical order, are as follows: Dr. Yoshihisa Fujino (present chairperson of the study group), Dr. Akira Ogami, Dr. Arisa Harada, Dr. Ayako Hino, Dr. Hajime Ando, Dr. Hisashi Eguchi, Dr. Kazunori Ikegami, Dr. Kei Tokutsu, Dr. Keiji Muramatsu, Dr. Koji Mori, Dr. Kosuke Mafune, Dr. Kyoko Kitagawa, Dr. Masako Nagata, Dr. Mayumi Tsuji, Ms. Ning Liu, Dr. Rie Tanaka, Dr. Ryutaro Matsugaki, Dr. Seiichiro Tateishi, Dr. Shinya Matsuda, Dr. Tomohiro Ishimaru, and Dr. Tomohisa Nagata. All members are affiliated with the University of Occupational and Environmental Health, Japan.

## Disclosure

### Ethical approval

This study conformed to the principles of the Declaration of Helsinki and was approved by the ethics committee of the University of Occupational and Environmental Health, Japan (reference No. R2-079 and R3-006).

### Informed Consent

Informed consent was obtained from all participants through the website. Registry and the Registration No. of the study/Trial: N/A

### Animal studies

N/A

